## Supplementary Materials for "Identification of genetic factors influencing metabolic dysregulation and retinal support for MacTel, a retinal disorder"

**Supplementary Material**

Supplementary Figures

**A** **B**

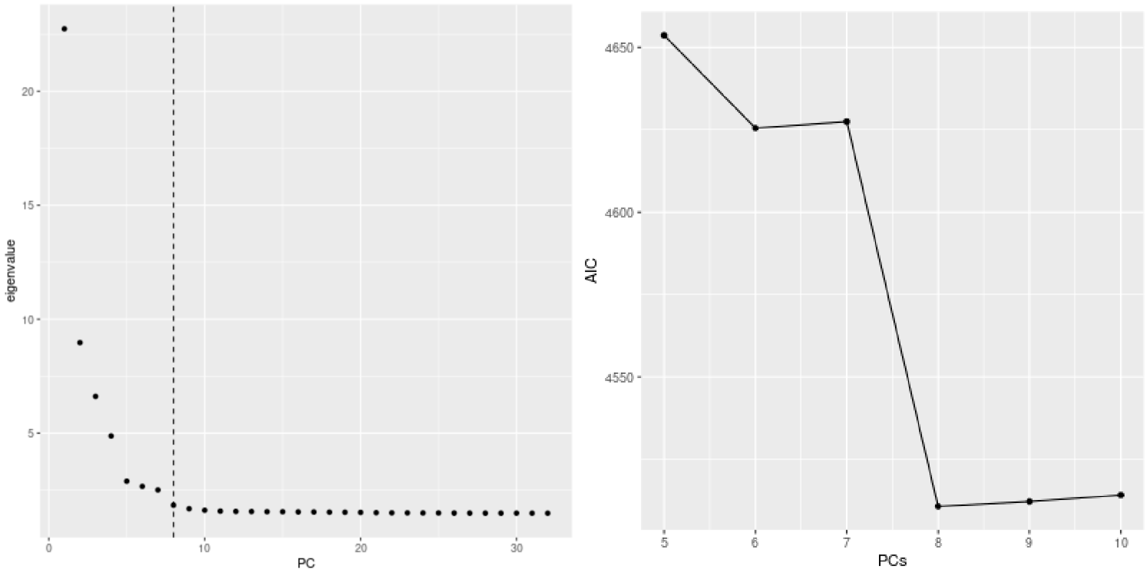


**Figure S1.** Selection of Principal Components (PCs) of ancestry to include as covariates in the GWAS. A) Scree Plot showing the eigenvalues of principal components 1 to 32. B) Plot showing the Akaike Information Criterion (AIC) for null models fitted with a varying numbers of principal components.

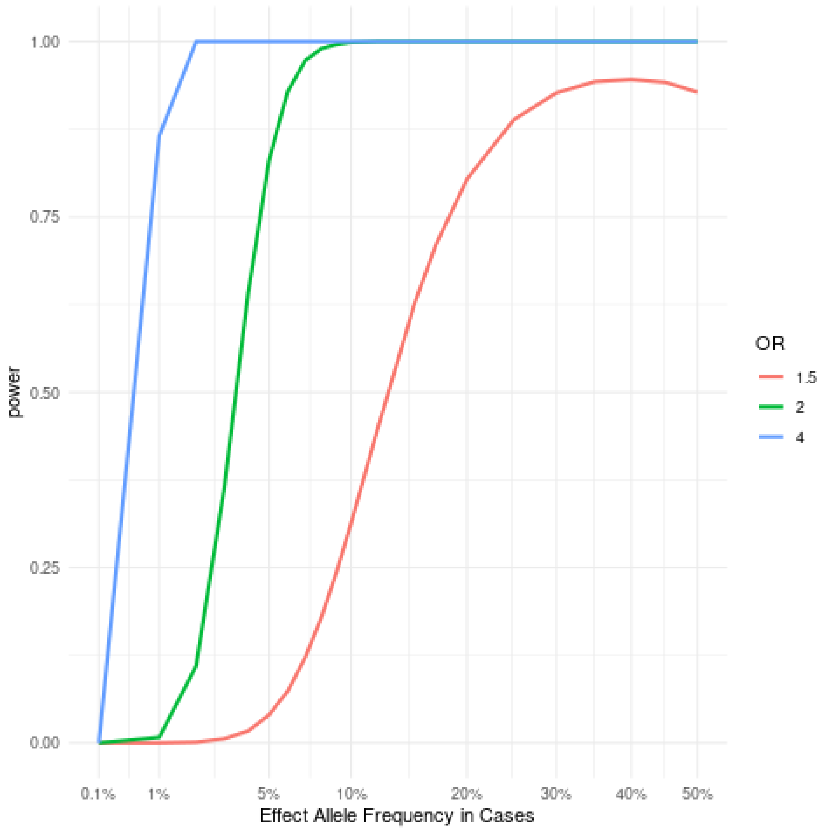


**Figure S2.** Plot of statistical power (y axis) vs effect allele frequencies (x axis), for detecting genome-wide significant associations in the present study (1067 cases vs 3799 controls), for varying ORs (colored lines). A population prevalence of MacTel of 0.45%, and an additive genetic model is assumed.

**A**

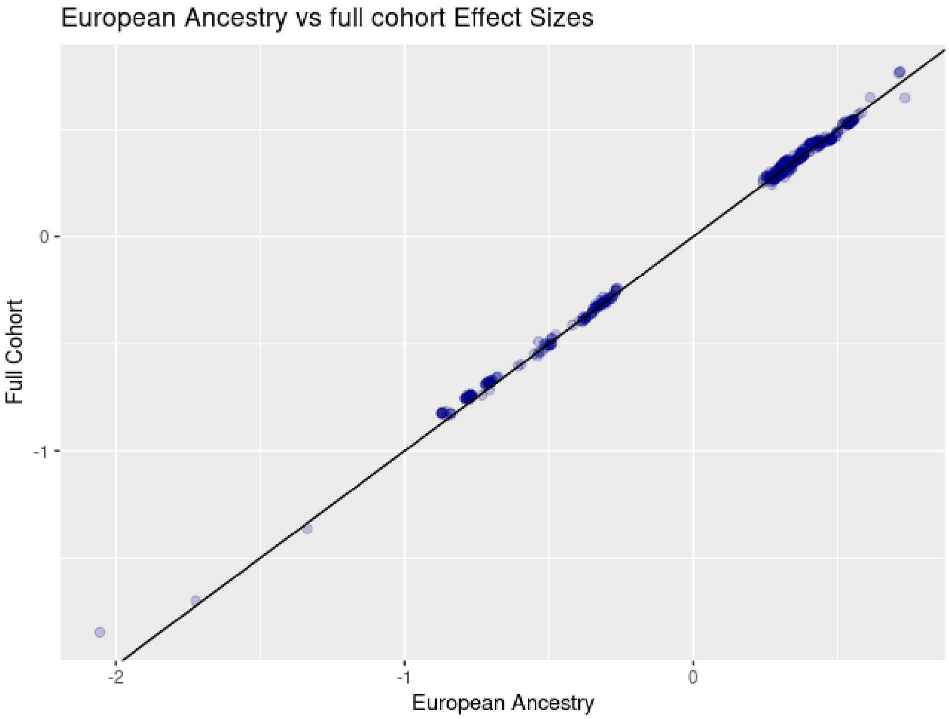


**B**

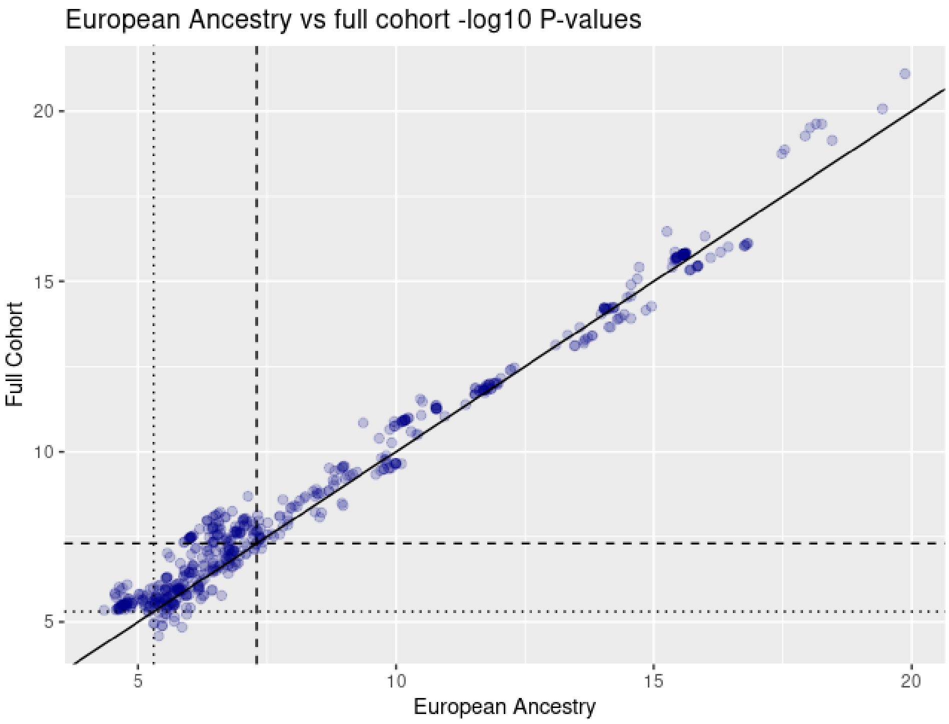


**Figure S3.** Comparison of A. effect sizes and B. P-values, from the full cohort analysis, and European Ancestry analysis. Shown are all SNPs meeting suggestive significance (P < 5E-6) in either analysis.

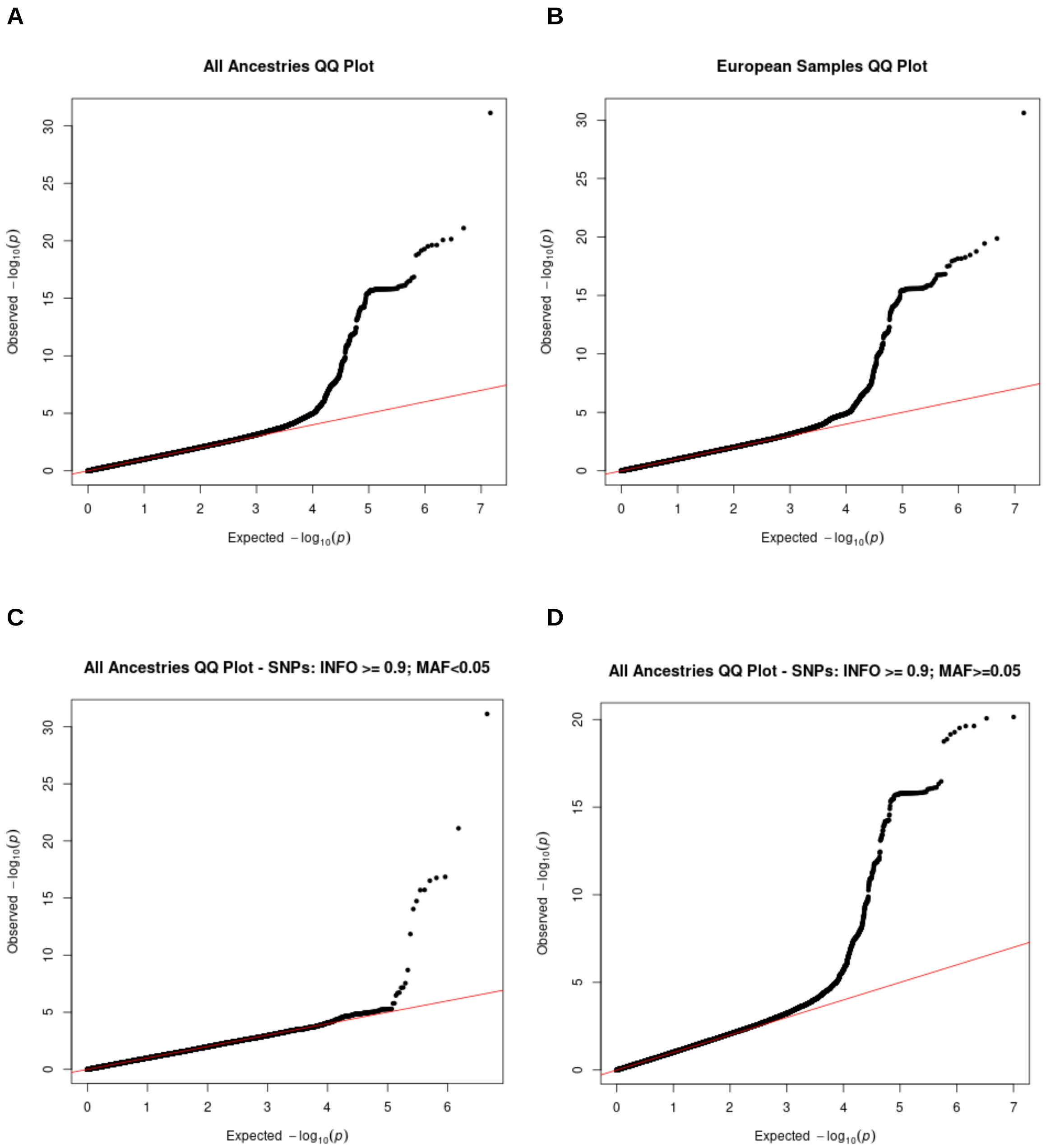
A
All Ancestries QQ Plot
B
European Samples QQ Plot
C
All Ancestries QQ Plot - SNPs: INFO >= 0.9; MAF<0.05
D
All Ancestries QQ Plot - SNPs: INFO >= 0.9; MAF>=0.05
Observed −log10(p)
Expected −log10(p)

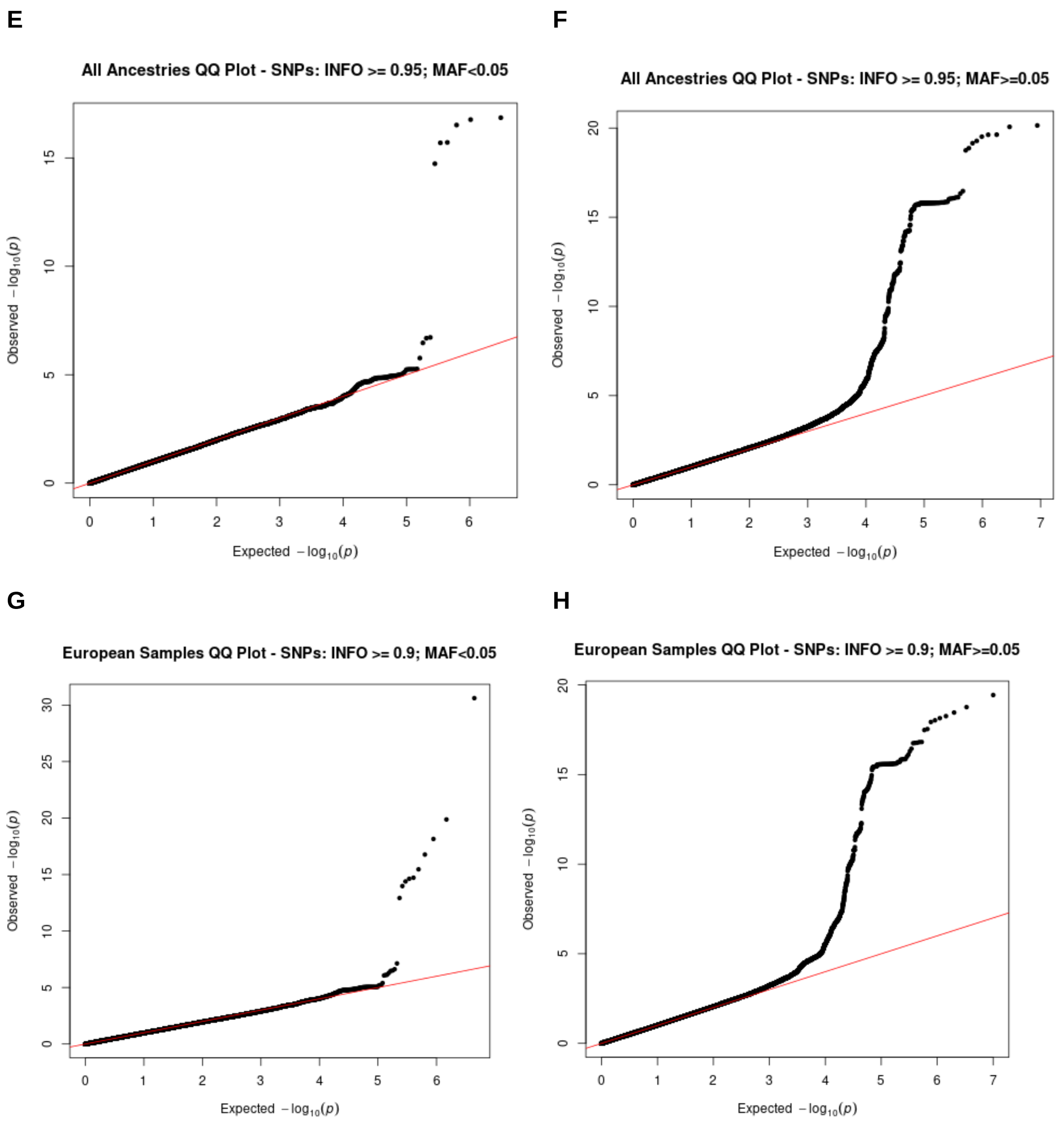
E
All Ancestries QQ Plot - SNPs: INFO >= 0.95; MAF<0.05
Observed $-\log_{10}(p)$
Expected $-\log_{10}(p)$
F
All Ancestries QQ Plot - SNPs: INFO >= 0.95; MAF>=0.05
Observed $-\log_{10}(p)$
Expected $-\log_{10}(p)$
G
European Samples QQ Plot - SNPs: INFO >= 0.9; MAF<0.05
Observed $-\log_{10}(p)$
Expected $-\log_{10}(p)$
H
European Samples QQ Plot - SNPs: INFO >= 0.9; MAF>=0.05
Observed $-\log_{10}(p)$
Expected $-\log_{10}(p)$

I

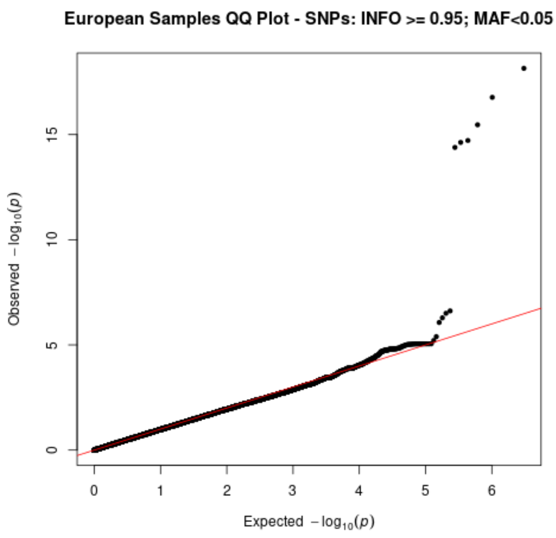


J

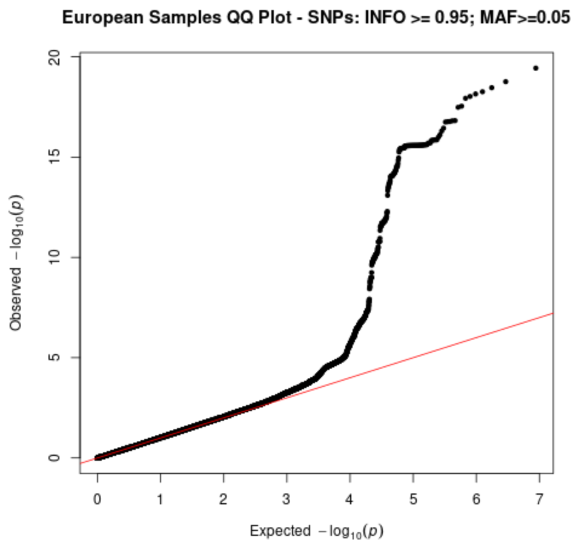


**Figure S4. Quantile-quantile plots**

A) All ancestry analysis - All SNPs (MAF>0.25% and INFO≥0.9) - $\lambda$=1.011

B) European Ancestry only analysis - All SNPs (MAF>0.25% and INFO≥0.9) - $\lambda$=1.013

C) All ancestry analysis - rare SNPs (MAF<5%), INFO≥0.9 - $\lambda$=0.992

D) All ancestry analysis - common SNPs (MAF≥5%) and INFO≥0.9 - $\lambda$=1.013

E) All ancestry analysis - rare SNPs (MAF<5%), INFO≥0.95 - $\lambda$=0.991

F) All ancestry analysis - common SNPs (MAF≥5%) and INFO≥0.95 - $\lambda$=1.014

G) European Ancestry analysis - rare SNPs (MAF<5%), INFO≥0.9 - $\lambda$=0.983

H) European Ancestry analysis - common SNPs (MAF≥5%) and INFO≥0.9 - $\lambda$=1.006

I) European Ancestry analysis - rare SNPs (MAF<5%), INFO≥0.95 - $\lambda$=0.984

J) European Ancestry analysis - common SNPs (MAF≥5%) and INFO≥0.95 - $\lambda$=1.009

**A**

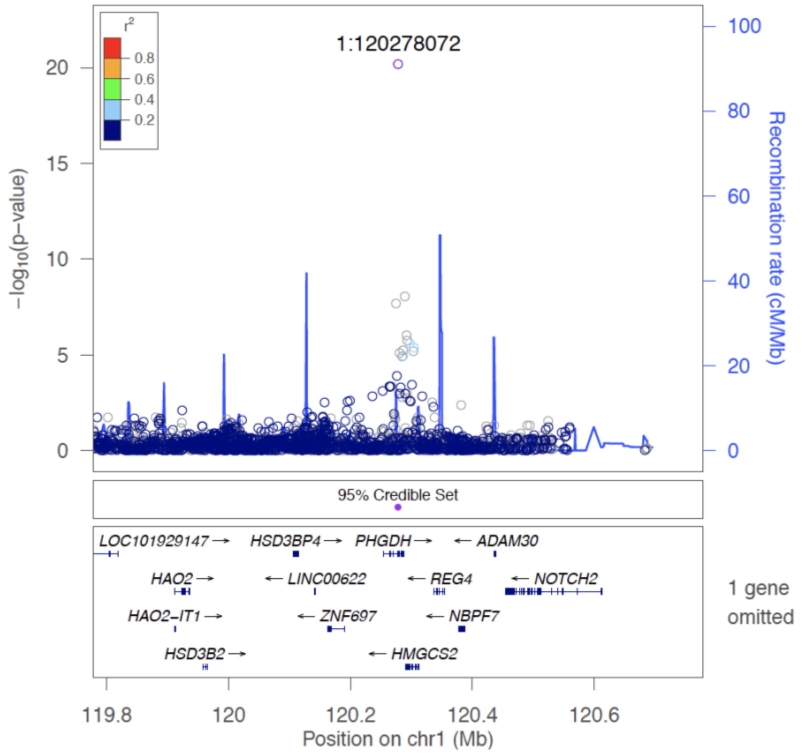
1:120278072
r²
0.8
0.6
0.4
0.2
20
15
10
5
0
−log10(p-value)
100
80
60
40
20
0
Recombination rate (cM/Mb)
95% Credible Set
LOC101929147
HSD3BP4
PHGDH
ADAM30
HAO2
LINC00622
REG4
NOTCH2
HAO2−IT1
ZNF697
NBPF7
HSD3B2
HMGCS2
1 gene omitted
119.8
120
120.2
120.4
120.6
Position on chr1 (Mb)

**B**

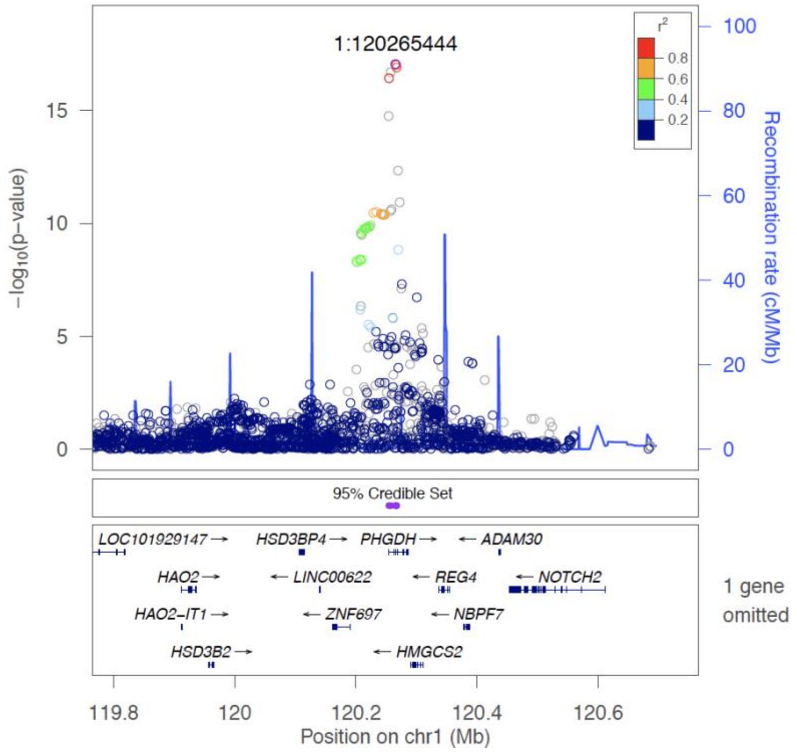
1:120265444
r²
0.8
0.6
0.4
0.2
15
10
5
0
−log10(p-value)
100
80
60
40
20
0
Recombination rate (cM/Mb)
95% Credible Set
LOC101929147
HSD3BP4
PHGDH
ADAM30
HAO2
LINC00622
REG4
NOTCH2
HAO2−IT1
ZNF697
NBPF7
HSD3B2
HMGCS2
1 gene omitted
119.8
120
120.2
120.4
120.6
Position on chr1 (Mb)

C

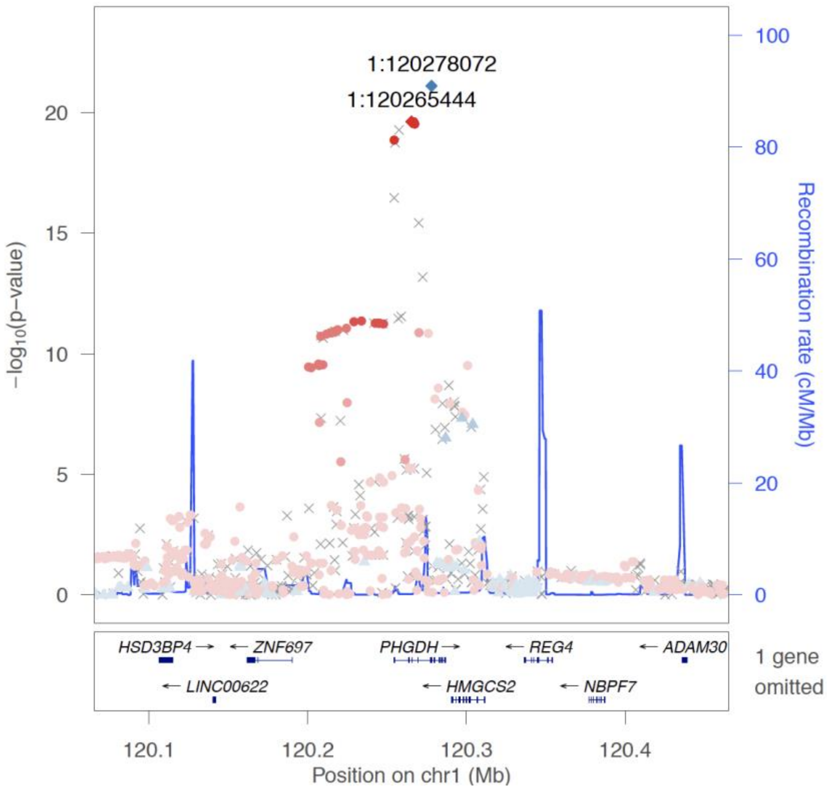
1:120278072
1:120265444
−log10(p−value)
Recombination rate (cM/Mb)
HSD3BP4 →
← ZNF697
PHGDH →
← REG4
← ADAM30
← LINC00622
← HMGCS2
← NBPF7
1 gene omitted
120.1
120.2
120.3
120.4
Position on chr1 (Mb)

D

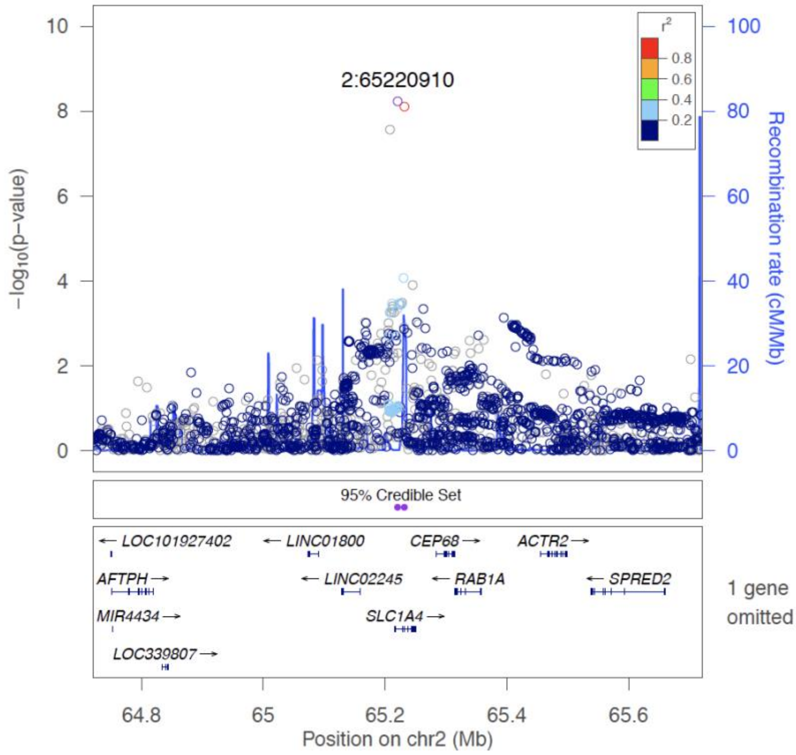
2:65220910
r²
0.8
0.6
0.4
0.2
−log10(p−value)
Recombination rate (cM/Mb)
95% Credible Set
← LOC101927402
← LINC01800
CEP68 →
ACTR2 →
AFTPH →
← LINC02245
← RAB1A
← SPRED2
MIR4434 →
SLC1A4 →
LOC339807 →
1 gene omitted
64.8
65
65.2
65.4
65.6
Position on chr2 (Mb)

E

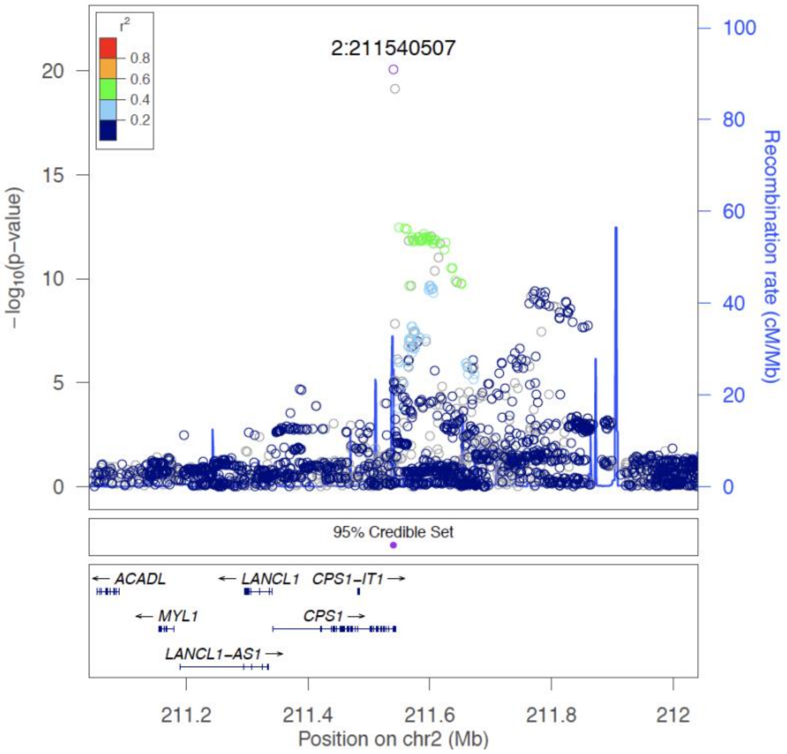

r²
0.8
0.6
0.4
0.2
2:211540507
−log10(p−value)
20
15
10
5
0
100
80
60
40
20
0
Recombination rate (cM/Mb)
95% Credible Set
← ACADL
← LANCL1
CPS1−IT1 →
← MYL1
CPS1 →
LANCL1−AS1 →
211.2
211.4
211.6
211.8
212
Position on chr2 (Mb)


F

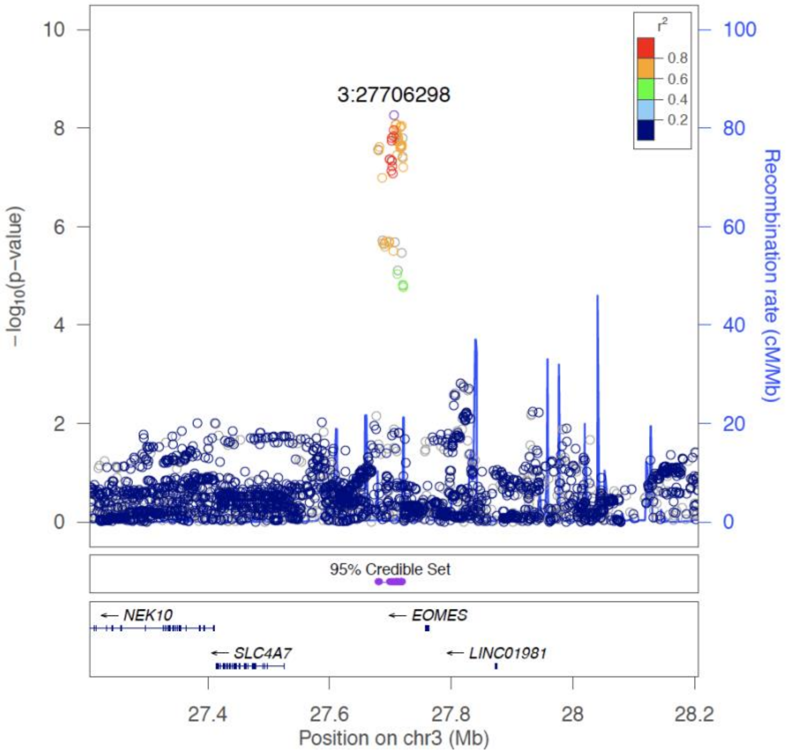

r²
0.8
0.6
0.4
0.2
3:27706298
−log10(p−value)
10
8
6
4
2
0
100
80
60
40
20
0
Recombination rate (cM/Mb)
95% Credible Set
← NEK10
← EOMES
← SLC4A7
← LINC01981
27.4
27.6
27.8
28
28.2
Position on chr3 (Mb)

G

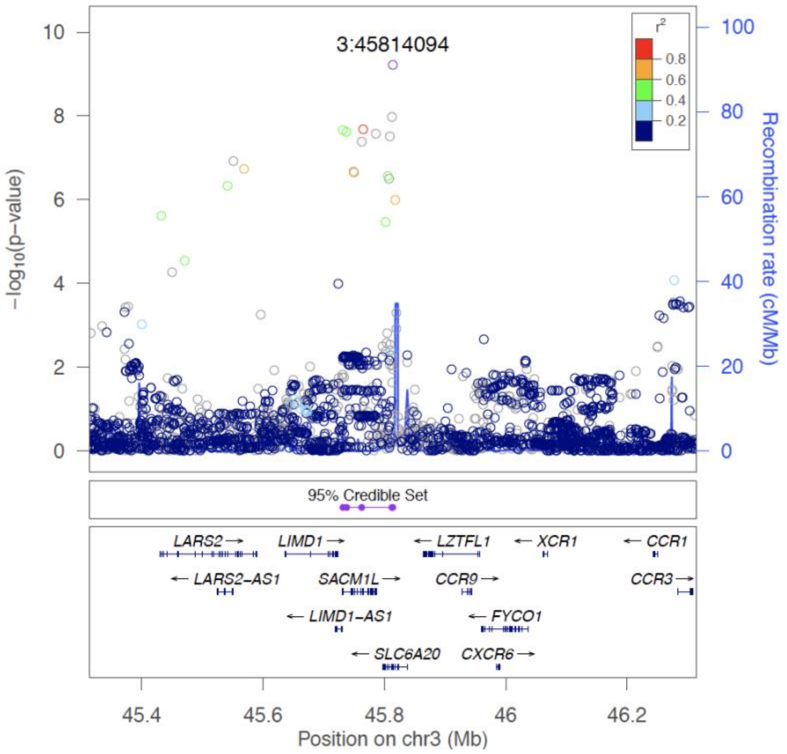

3:45814094
r²
0.8
0.6
0.4
0.2
−log10(p−value)
Recombination rate (cM/Mb)
95% Credible Set
LARS2
LIMD1
LZTFL1
XCR1
CCR1
LARS2−AS1
SACM1L
CCR9
CCR3
LIMD1−AS1
FYCO1
SLC6A20
CXCR6
45.4
45.6
45.8
46
46.2
Position on chr3 (Mb)


H

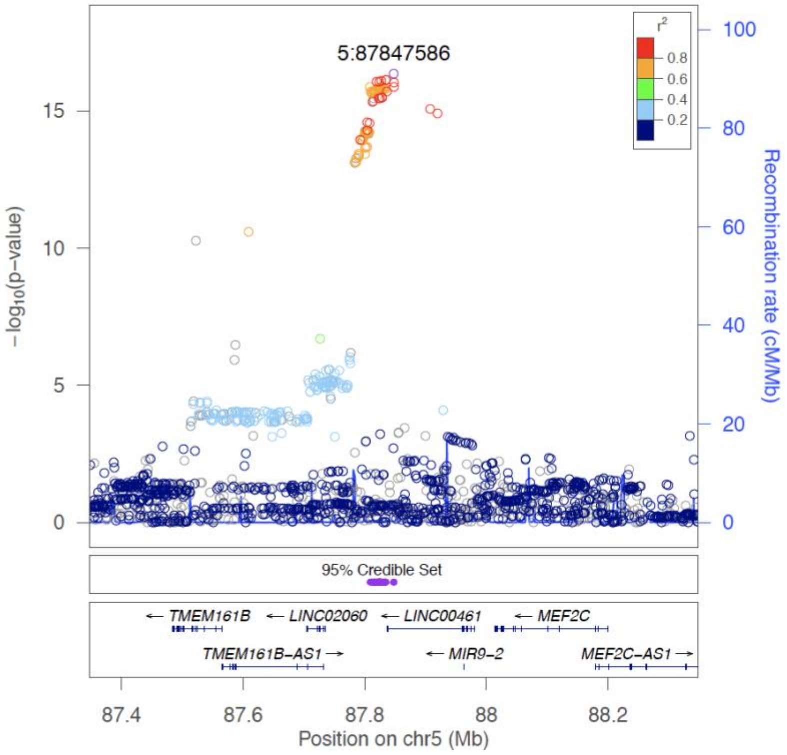

5:87847586
r²
0.8
0.6
0.4
0.2
−log10(p−value)
Recombination rate (cM/Mb)
95% Credible Set
TMEM161B
LINC02060
LINC00461
MEF2C
TMEM161B−AS1
MIR9−2
MEF2C−AS1
87.4
87.6
87.8
88
88.2
Position on chr5 (Mb)

I

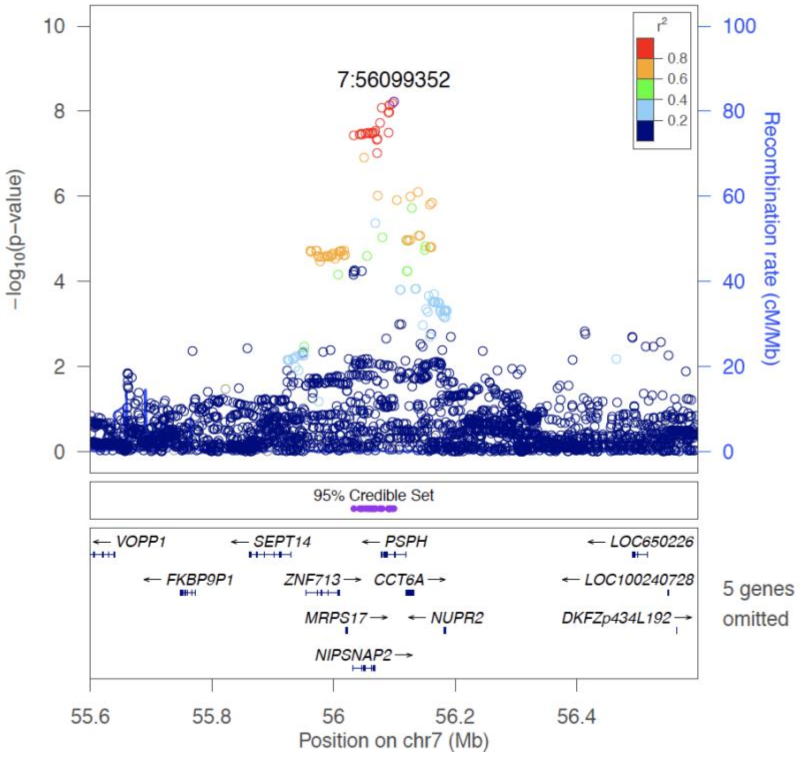
7:56099352
−log10(p−value)
Recombination rate (cM/Mb)
r2
95% Credible Set
VOPP1
SEPT14
PSPH
LOC650226
FKBP9P1
ZNF713
CCT6A
LOC100240728
MRPS17
NUPR2
DKFZp434L192
NIPSNAP2
5 genes omitted
Position on chr7 (Mb)

J

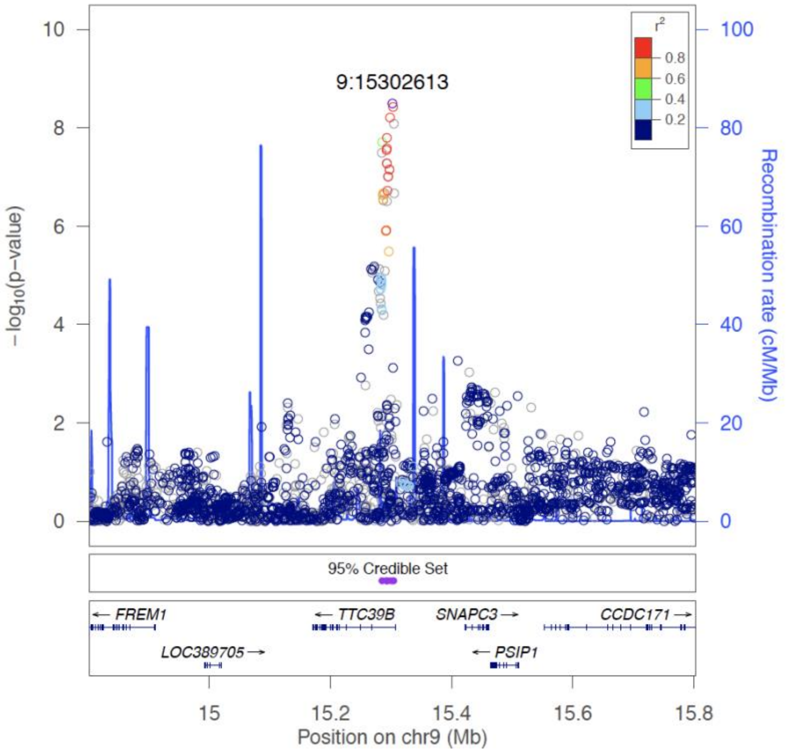
9:15302613
−log10(p−value)
Recombination rate (cM/Mb)
r2
95% Credible Set
FREM1
TTC39B
SNAPC3
CCDC171
LOC389705
PSIP1
Position on chr9 (Mb)

K

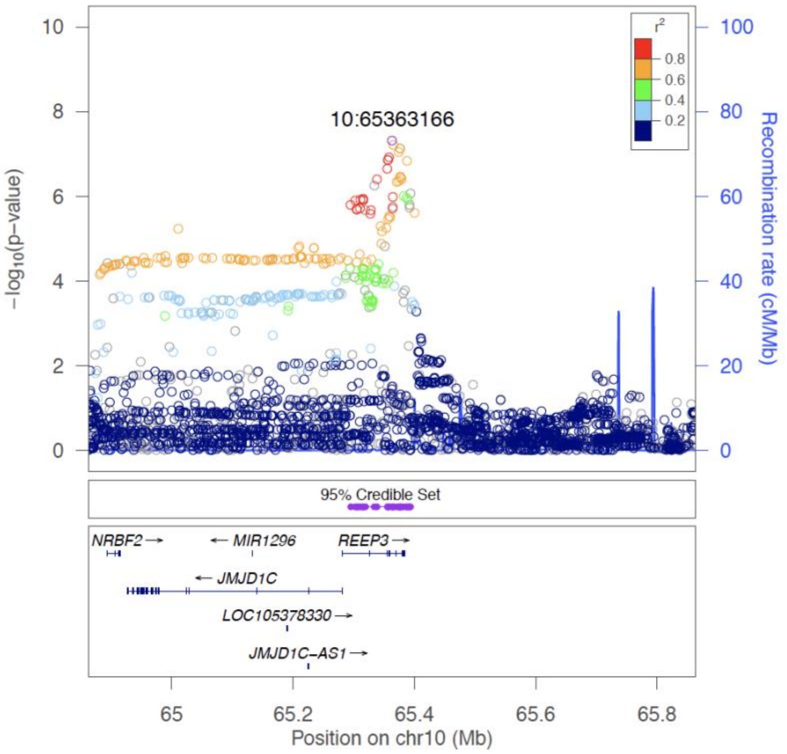
10:65363166
r²
0.8
0.6
0.4
0.2
−log10(p−value)
Recombination rate (cM/Mb)
95% Credible Set
NRBF2
MIR1296
REEP3
JMJD1C
LOC105378330
JMJD1C−AS1
65
65.2
65.4
65.6
65.8
Position on chr10 (Mb)

L

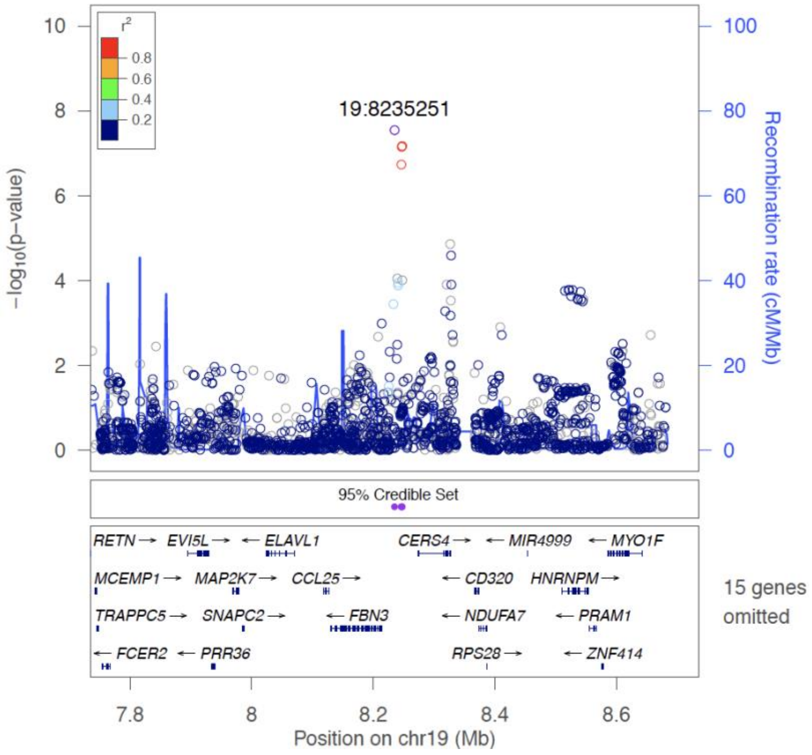
19:8235251
r²
0.8
0.6
0.4
0.2
−log10(p−value)
Recombination rate (cM/Mb)
95% Credible Set
RETN
EVI5L
ELAVL1
CERS4
MIR4999
MYO1F
MCEMP1
MAP2K7
CCL25
CD320
HNRNPM
TRAPPC5
SNAPC2
FBN3
NDUFA7
PRAM1
FCER2
PRR36
RPS28
ZNF414
15 genes omitted
7.8
8
8.2
8.4
8.6
Position on chr19 (Mb)

**Figure S5.** LocusZoom Plots for GW-significant loci including 95% CI for credible SNP set (purple bars). Gene boundaries are displayed by blue bars below plot.
A) 1p12 signal 1: Top SNP rs146953046 (chr1:120278072) - P-values conditional on rs146953046 (chr1:120265444).
B) 1p12 signal 2: Top SNP rs146953046 (chr1:120265444) - P-values conditional on rs146953046 (chr1:120278072).
C) 1p12 region with two independent signals: rs146953046 (chr1:120278072) and rs146953046 (chr1:120278072). Non-conditional P-values shown. SNPs in LD with rs146953046 (chr1:120278072) coloured in blue; SNPs in LD with rs146953046 (chr1:120265444) coloured in red.
D) 2p14 locus: Top SNP rs2160387 (chr2: 65220910).
E) 2q34 locus: Top SNP rs1047891 (chr2:211540507).
F) 3p24.1 locus: Top SNP rs9820465 (chr3:27706298).
G) 3p21.31locus: Top SNP rs17279437 (chr3:45814094).
H) 5q14.3 locus: Top SNP rs17421627 (chr5:87847586). I
) 7p11.2 locus: Top SNP rs6955423 (chr7:56099352).
J) 9p22.3 locus: Top SNP rs677622 (chr9:15302613).
K) 10q21.3 locus: Top SNP rs10995566 (chr10:65363166).
L) 19p13.2 locus: Top SNP rs139412173 (chr19:8235251).

**A**

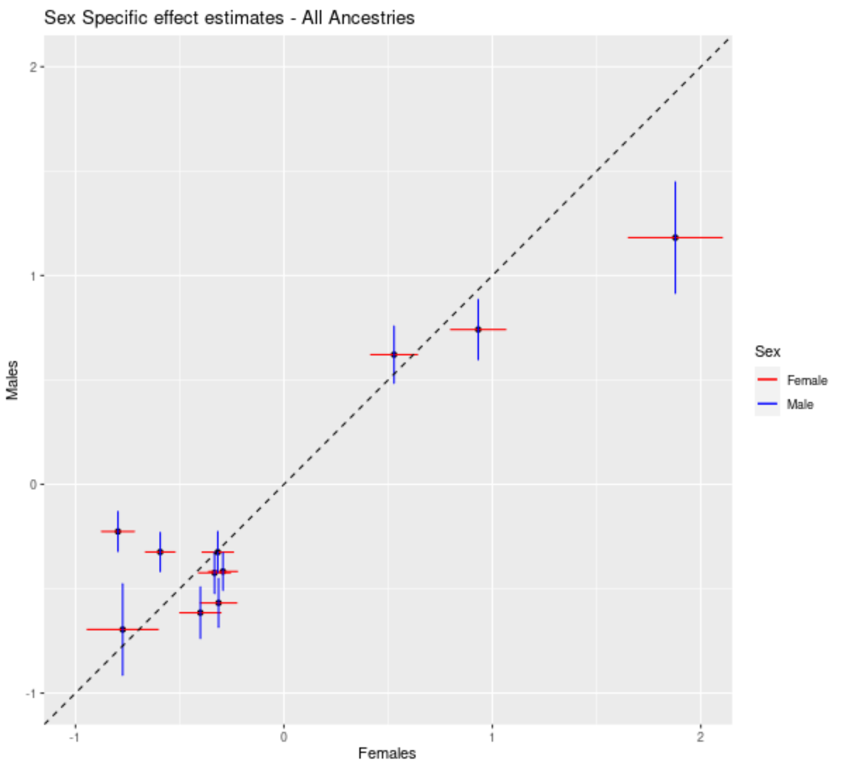


**B**

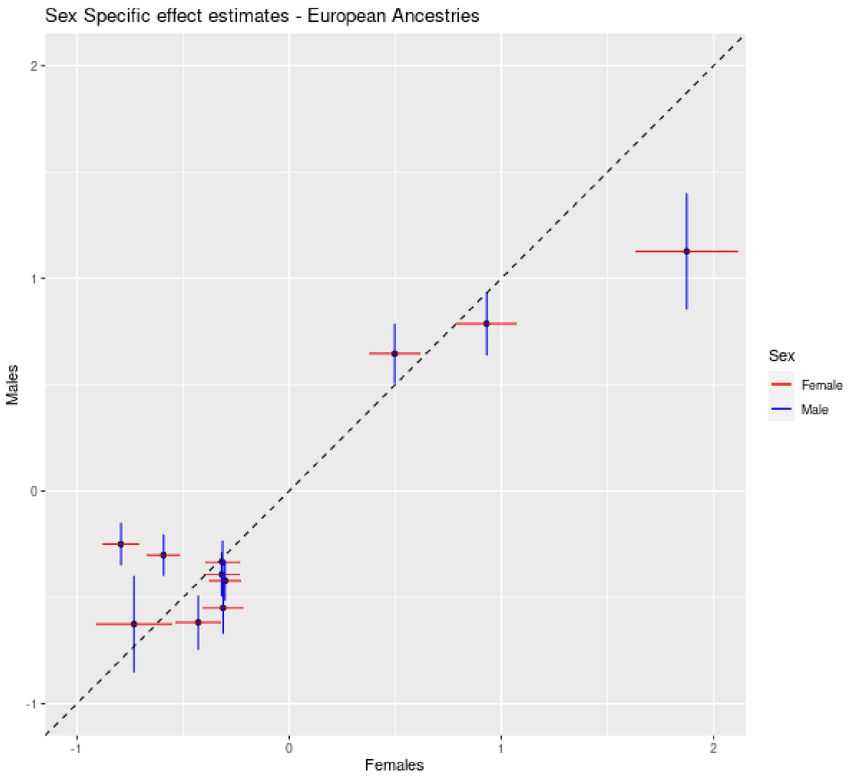


**Figure S6.** Comparison of effect estimates (log odds ratios) in females (x-axis) and males (y-axis). Red and blue bars indicate standard errors of effect estimates. A) All ancestries analysis. B) European Ancestries analysis.

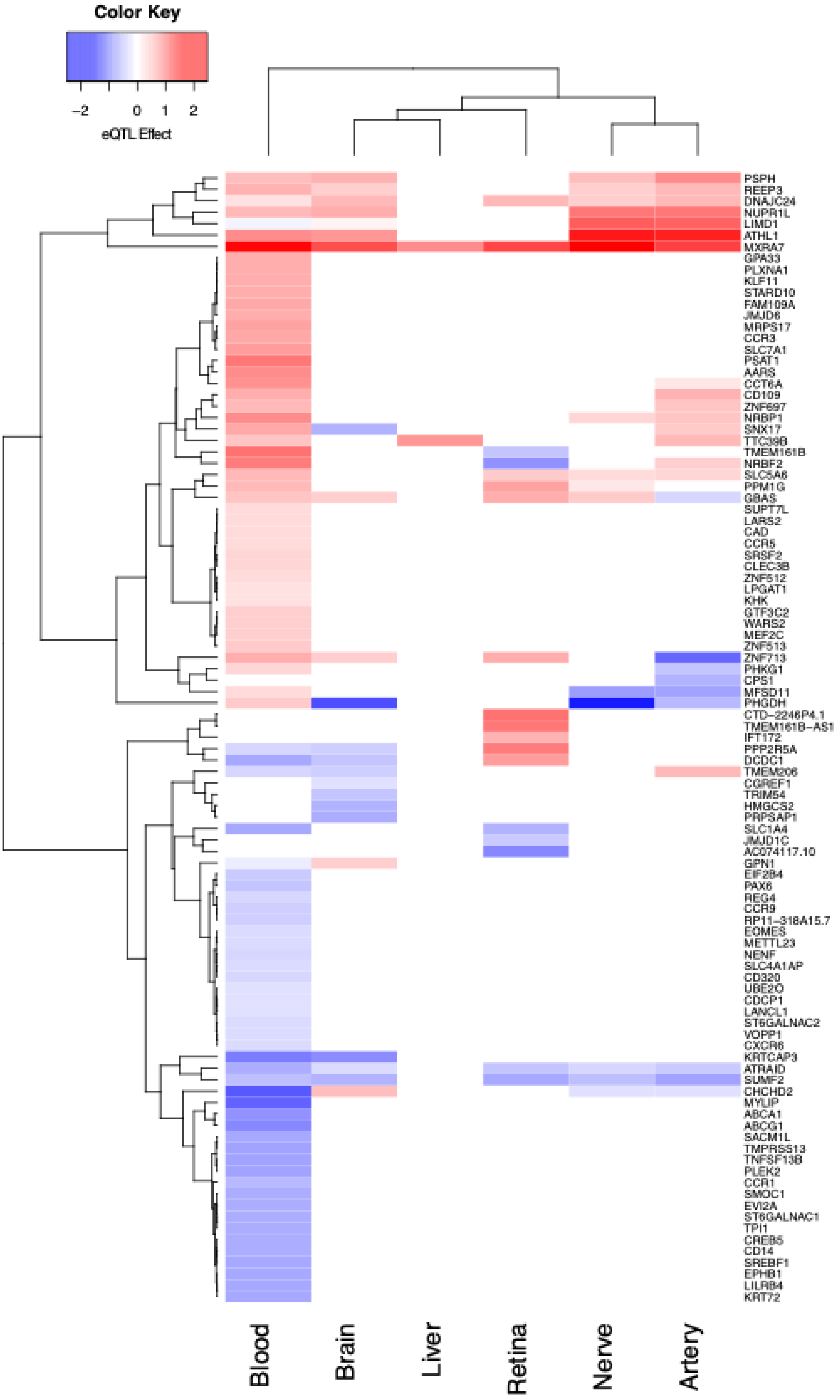


**Figure S7. eQTL effects heatmap.** The heatmap presents eQTL effects found across the analysed tissues. The average effect size of eQTL for each gene in each tissue is presented in the heatmap, non-significant eQTL are displayed in white. Genes and tissues are ordered according to hierarchical clustering modelling as presented in the image.

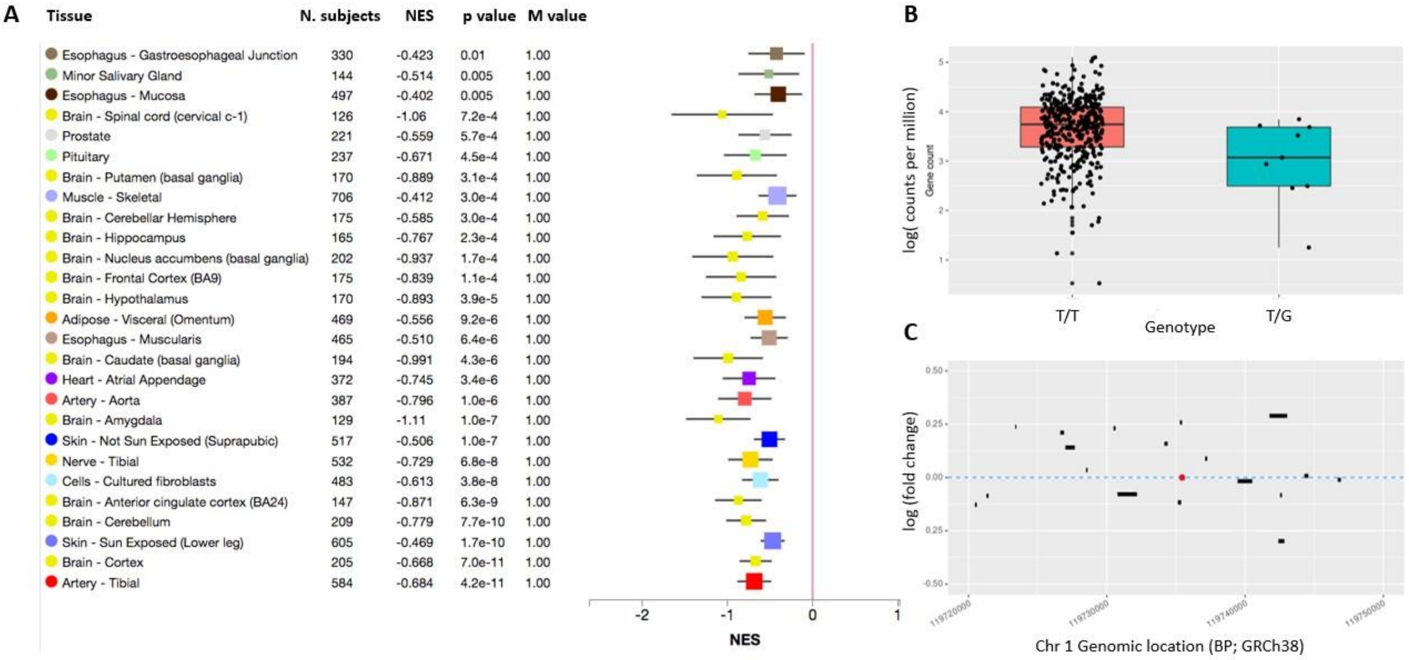


**Figure S8. Effects of the rare deleterious SNP rs146953046 on gene expression and exon abundance in the GTEx database and the retina. A)** Tissues in the GTEx database (y axis) for which the minor (G) allele of rs146953046 is associated with a significant difference in *PHGDH* gene expression with high confidence (M value =1). NES: normalized effect size (equivalent to alternative SNP beta value in a linear model); M value: posterior probability of eQTL. Horizontal whisker plots indicate the median (filled square) and 95% confidence interval (black bars) for NES estimates. In all tissues the effect of the G allele is suppressive. **B)** Expression of *PHGDH* in the retina (y axis; Ratnapriya *et al* 2019, *Nat Genet*) for individuals with the reference genotype (T/T) compared to heterozygotes (x axis). Each point represents *phgdh* expression measured in an individual donor retina. Box plot centre line: median; box limits: upper and lower quartiles; whiskers: 1.5x interquartile range. The difference between genotypes is significant ($p < 0.003$) after correcting for subject age, sex and AMD status. **C)** Relative expression of merged *PHGDH* exons in the neural retina between heterozygous individuals and reference homozygous individuals. The span of each exon across chromosome 1 (x axis) is indicated by black bars. The height of the exon bars relative to the y axis indicates log fold change. The red point indicates the genomic (intronic) location of the rare SNP.

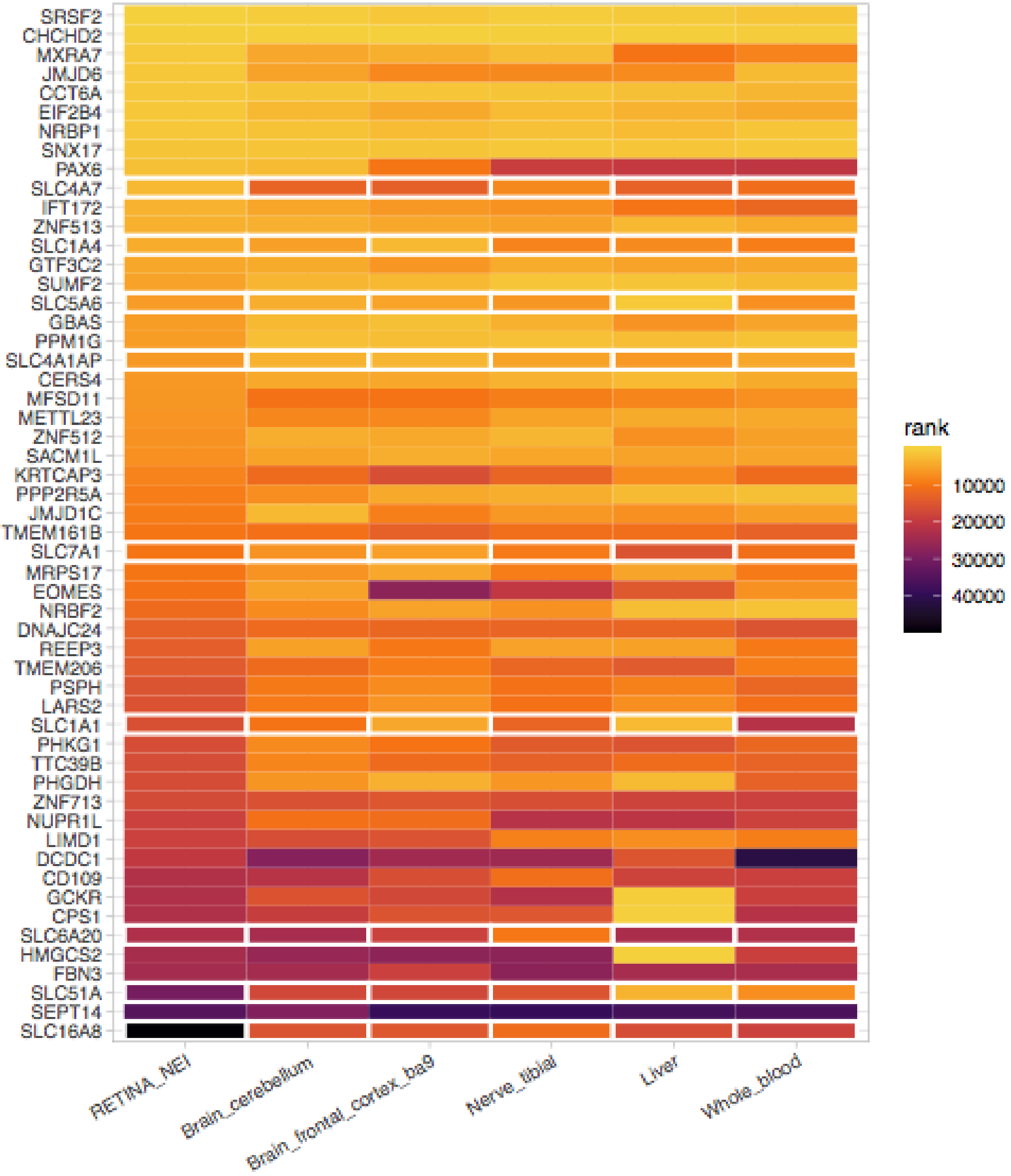


**Figure S9. Ranked length-normalized gene expression (transcripts per million) for 55 genes prioritized in post-GWAS analyses, across multiple human tissues.** Tiles outlined in white are solute-transport family genes. Tile color indicates transcriptional abundance rank within tissue (1 = highest expression rank = yellow). Median TPM for retina calculated based on gene expression data from 106 healthy retina donors, published by Ratnapriya et al. Expression for other tissues taken from GTEx supplementary data.

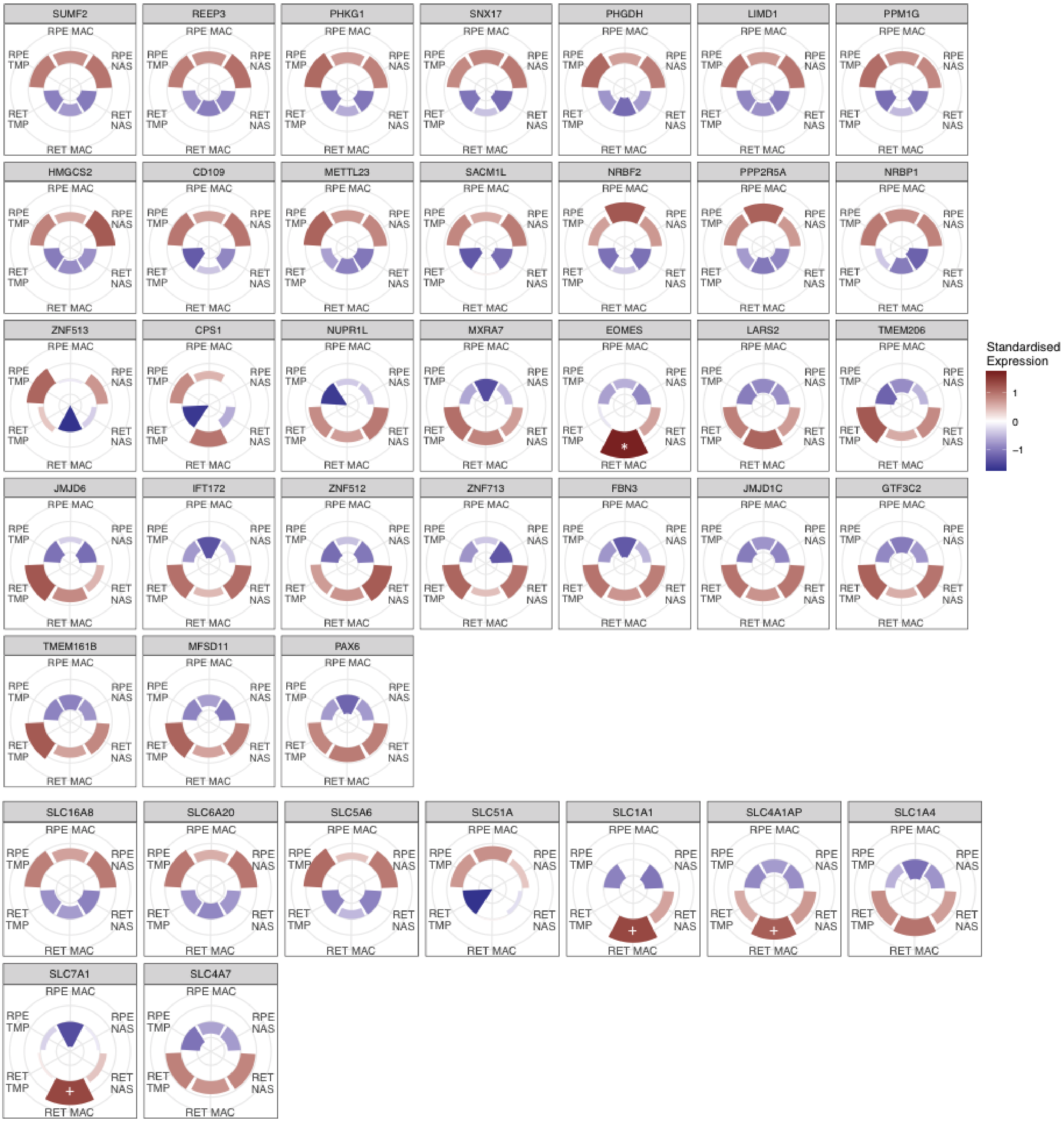


**Figure S10.** Relative expression patterns across retina (RET) and retinal pigment epithelium (RPE) for genes displaying significant differences between the two tissues, and between retinal regions. SLC family genes are grouped at the bottom (see also Table S12). TMP: temporal side; NAS: nasal side; MAC: macular region. NB SLC1A1 is not differentially abundant between RET and RPE, but shows area-specific enrichment. * enriched in MAC relative to NAS and TMP; + enriched in MAC relative to TMP.

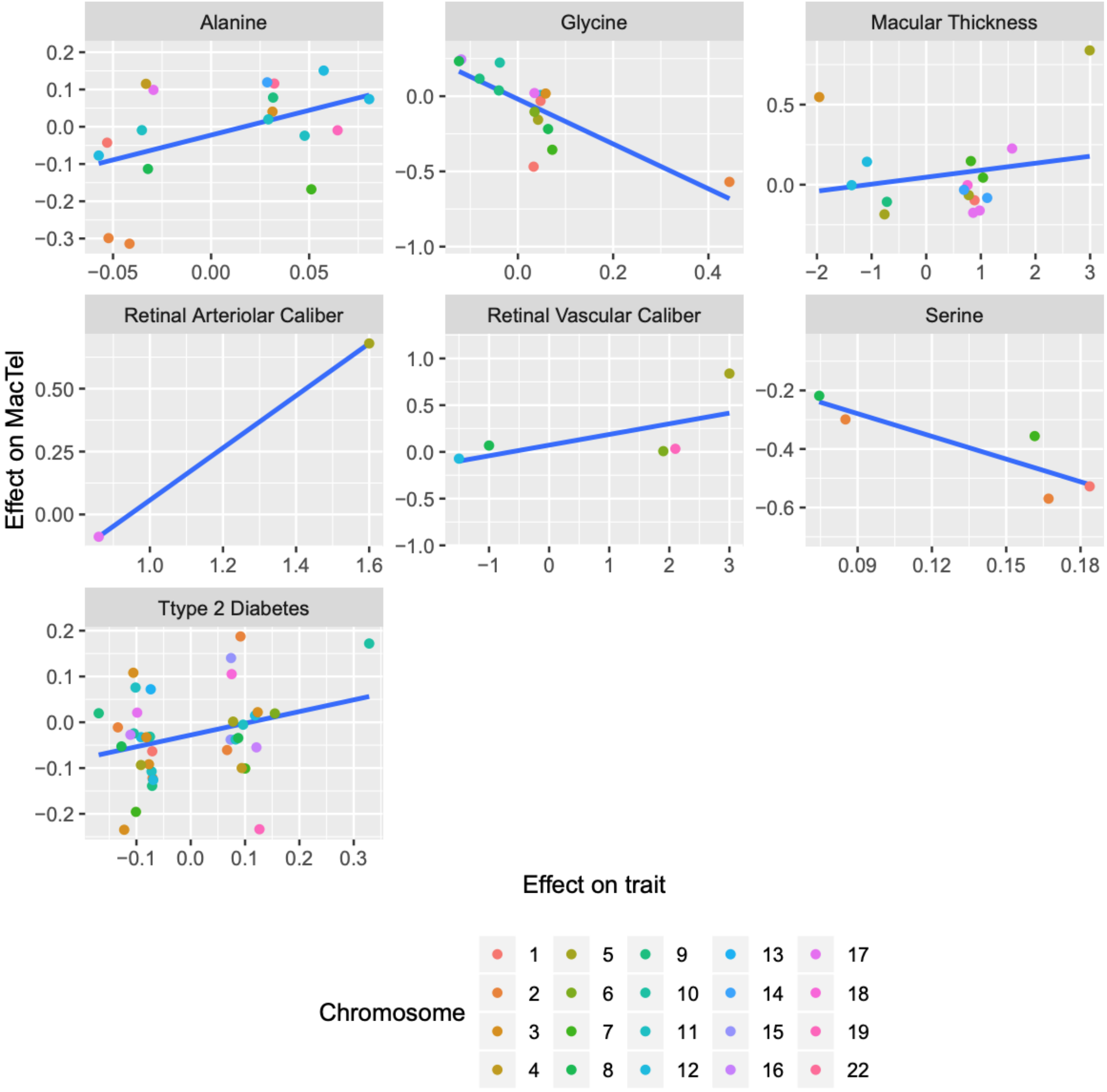


**Figure S11.** MacTel vs phenotype effect size comparison of SNPs instrument used in Mendelian Randomization. Each panel contains the effect size comparison for SNPs used in each phenotype. Each SNP is presented as a point, and different colours represent different chromosomes. X-axis presents the effect size on each phenotype of interest while y-axis presents the effect on MacTel disease.

**Supplementary Table Legends**

Table S1. Summary of SNP and Sample-based quality control:
A. Tranche 1 - MacTel consortium samples genotyped on Omni5-exome (pre-imputation);
B. Tranche 2 - AREDs controls genotyped on Omni-2.5 (pre-imputation);
C. Tranche 3 - MacTel Consortium and Twinning samples genotyped on the Global Screening Array (pre-imputation);
D. All tranches merged (post-imputation).

Table S2. Sample composition and genotyping platform for cases and controls included in the full cohort, and European-only sub-set.

Table S3. 95% credible sets, of causal SNPs at each GW-significant locus, as determined by Bayesian fine-mapping.[1]

Table S4. Haplotype frequencies for the two *PHGDH* SNPs (rs146953046 and rs532303), in cases and controls. The alleles associated with increased MacTel risk are the G allele for rs146953046 and the A allele for rs532303.

Table S5. Sex Interaction Analysis. Association results for the 11 genome-wide significant SNPs, in males and females separately. For each SNP, sex-interaction was formally tested using Welch's t-test.

Table S6. MAGMA results tables as performed by FUMA. Only genes with FDR<0.05 are shown. NSNPs is the number of SNPs used by MAGMA to calculate principal components for each gene. The number of PCs used for each gene is presented in NPARAM.

Table S7. List of parameters used in FUMA. eQTL tissues can be found in parameter "eqtlMaptss".

Table S8. Summary of eQTL results table. This table presents the average effect of all MacTel risk SNPs affecting gene expression for each gene in each tissue. nSNP is the significant eSNPs for each gene in each tissue. nTissue is the count of tissue presenting significant eSNPs for each gene. Global mean effect is the average among tissue specific effects. Direction indicated whether the gene should be positively or negatively regulated by the SNPs increasing MacTel risk.

Table S9. Genetic colocalization results for MacTel genome-wide significant loci compared to retinal eQTL loci and GW-significant loci for MacTel-related traits. Candidate gene: gene bounding, or closest to MacTel GWAS locus; N. SNPs: number of SNPs represented in both GWAS / eQTL result sets used for colocalization analysis; PP shared causal variants: posterior probability that the Mactel GWAS and Retina eQTL/related trait GWAS signals derive from a shared variant; PP distinct causal variants: Posterior probability that the Mactel GWAS and Retina eQTL/related trait GWAS signals derive from distinct variants. P-values in bold meet the gene-specific threshold for a significant eQTL as determined by Ratnapriya et al [2]. Posterior probabilities for shared causal variants >0.75 (bold text) are considered strong evidence of genetic colocalization.

Table S10. List of significant and suggestive-significant genes for each tissue from the TWAS.

Table S11. List of prioritised genes by any analysis. GW_sig_snp is 1 if the gene was located underneath a locus reaching GW significance. Sugg_GW_sig_snp is 1 if the gene was located underneath a locus reaching GW significance or suggestive significance. eQTL is 1 if the gene was significantly affected in any tissue by any SNPs in LD (r2>0.4) with significant or suggestive significant loci. MAGMA is 1 if the gene was significant in the MAGMA analysis. TWAS is 1 if the gene was significant in the TWAS analysis. N_evidence is the sum of all evidence.

Table S12. Retina vs RPE tissue expression comparison results based on Whitmore et al 2014 *Exp Eye Res*. Additionally, comparison between nasal (NAS), temporal (TMP) and macular (MAC) regions of the retina/RPE is presented for all genes.

Table S13. Single cell expression comparison results based on three studies detailed in Online Methods. The cell type exhibiting maximal expression for each gene is presented in cell_max_expression. Average increase of expression between that cell type and all other cell types is represented in the Effect column.

Table S14. LD-score regression results from LDhub database and an additional 13 ocular phenotypes not included in LDhub. Traits with a significant genetic correlation with MacTel are those with p-values below 0.05. Reference study details are provided in online methods. LDhub databases and studies contain subjects of predominantly European ancestry. SNPs from the major histocompatibility complex region (chr6 26M~34M) were excluded from this analysis. SE: standard error; Z: z score statistics for correlation significance.

Table S15. Mendelian Randomization results. Each tested metabolite PRS is presented in columns Metabolite. Effect of genetically predicted metabolic abundance on disease risk is presented in column effect and OR.

Table S16. MacTel Consortium members and affiliations.

**Supplementary Note**

Recruiting institutes and ethics approvals

The MacTel Project consortium recruited cases and controls at 24 participating clinical centers in seven countries (Australia, Germany, France, UK, Switzerland, Israel and United States). Informed written consent was obtained in accordance with ethics protocols for human subjects approved by the appropriate governing body at each site in accordance with the Declaration of Helsinki. Protocols and records of consent were centrally managed by the EMMES Corporation. The following ethics boards granted approval for human subject enrollment: Quinze-Vingts, Paris, France: Comité de Protection des Personnes Hôpital Saint-Antonie; Centre for Eye Research, Victoria, Australia: The Royal Victorian Eye and Ear Hospital; QIMR Berghofer Institute of Medical Research, Queensland, Australia; Clinique Ophtalmolgie de Creteil, Paris, France: Comité de Protection des Personnes Hôpital Saint-Antonie; Hospital Lariboisiere, Paris, France: Comité de Protection des Personnes Hôpital Saint-Antonie; Jules Stein Eye Institute, UCLA, California, USA: The UCLA Institutional Review Board; Lions Eye Institute, Nedlands, Australia: Sire Charles Gairdner Group Human Research Ethics Committee; Manhattan Eye, Ear and Throat Hospital, New York, USA: Lenox Hill Hospital Institutional Review Board; Moorfields Eye Hospital, London, UK: National Research Ethics Service; Retina Associates of Cleveland, Inc., Cleveland, Ohio, USA: Sterling Institutional Review Board; Save Sight Institute, Sydney, Australia: South Eastern Sydney Illawarra Area Health Service Human Research Ethics Committee–Northern Hospital Network; Scripps Research Institute, La Jolla, California, USA: Scripps Institutional Review Board; St. Franziskus Hospital, Munster, Germany: Ethik-Kommission der Arztekammer Westfalen-Lippe und der Medizinishchen Fakultat der Westfallschen Wilhelms-Universitat; The Goldschleger Eye Institute, Tel Hashomer, Israel: Ethics Committee The Chaim Sheba Medical Center; The New York Eye and Ear Infirmary, New York, USA: The Institutional Review Board of the New York Eye and Ear Infirmary; The Retina Group of Washington, Olympia, Washington, USA: Western Institutional Review Board; University of Bonn, Bonn, Germany: Rheinische Friedrich-Wilhelms- Universität Ethik-Kommission; University of Chicago, Chicago, Illinois, USA: The University of Chicago Division of Biological Sciences–The Pritzker School Institutional Review Board; University of Michigan, Ann Arbor, Michigan, USA: Medical School Institutional Review Board (IRBMED); University of Wisconsin, Madison, Wisconsin, USA: Office of Clinical Trials University of Wisconsin School of Medicine and Public Health; The Wilmer Eye Institute of Johns Hopkins University, Baltimore, Maryland, USA: Johns Hopkins School of Medicine Office of Human Subjects Research; Scheie Eye Institute University of Pennsylvania, Philadelphia, Pennsylvania, USA: University of Pennsylvania Office of Regulatory Affairs; University of Bern, Bern, Switzerland: Kantonale Ethikkommission Bern; John Moran Eye University of Utah, Salt Lake City, Utah, USA: The University of Utah Institutional Review Board; Bascom Palmer Eye Institute University of Miami, Miami, Florida, USA: The University of Miami Human Subjects Research Office; Columbia University, New York, New York, USA: Columbia University Medical Center Institutional Review Board Category 4 waiver for research involving specimens obtained from deidentified subjects.

**Supplementary Methods**

Patient Diagnosis

Protocol details of the multi-center Natural History Study of Macular Telangiectasia (MacTel Study) have been reported previously [3,4]. The diagnosis of MacTel type 2 was based on characteristic morphologic findings on fundoscopy, optical coherence tomography (OCT), fundus fluorescein angiography (FFA) and fundus autofluorescence, and was confirmed by the Moorfields Eye Hospital Reading Centre, London, UK.

### Sex Interaction analysis

For the top SNPs at each identified locus, we undertook analyses in males and females separately, with adjustment for 8 PCs of ancestry. In the all ancestries analyses, the females comprised of 644 cases and 2974 controls, and the males comprised of 423 cases and 825 controls. In the European ancestries analyses, the females comprised of 544 cases and 2941 controls, and the males comprised of 387 cases and 804 controls.
Given the unequal numbers of males and females, we formally tested for a sex interaction using Welsch's t-test:

$$t = \frac{\beta_F - \beta_M}{\sqrt{se_f^2 + se_M^2}}$$

With degrees of freedom:

$$d.f. = \frac{(se_F^2 + se_M^2)^2}{\frac{se_F^2}{n_f - 1} + \frac{se_M^2}{n_M - 1}}$$

P-values < $4.5 \times 10^{-3}$ (Bonferroni correction for 11 SNPs), were considered significant evidence of a sex interaction.

Polygenic Risk Scores

We employed the software PRSice2 [2-4] to construct a third, even less stringent PRS comprising a much larger number of SNPs. Given the rarity of MacTel, we were not able to source an independent dataset to validate the prediction power of the PRS created by PRSice. In lieu of this, we tested the prediction model within the discovery dataset. We observed the PRS with the highest $r^2$ to contain 34,424 SNPs. This model was additionally different from the PRS with the lowest empirical p-value (613 SNPs) thus likely overfitting the model. For this reason and the lack of a truly independent validation set, we decided to discard the PRS estimated using PRSice.

**Supplementary Results**

Inflation of test statistics

QQ plots (Figure S4) and assessment of the genomic control inflation factors ($\lambda_{GC}$), indicated that population stratification was appropriately accounted for in both analyses (European analysis: $\lambda_{GC}$=1.013; full-cohort analysis: $\lambda_{GC}$=1.011). We additionally used LD score regression (LDSC) to assess confounding due to population stratification, however the LDSC intercepts were less than one in both analyses (European analysis: intercept = 0.974; full-cohort analysis: intercept = 0.975), likely due to the small sample size.

Mendelian Randomization

Consistent with the previously published results, we found type 2 diabetes PRS to be robustly associated with disease risk, this result remained significant even after inclusion of genetically-predicted serine, glycine and alanine in the model. MR-Egger test resulted non-significant for all three metabolites as well as T2D ($b_0$Ser P=0.76; $b_0$Gly P=0.86; $b_0$Ala P=0.40; $b_0$T2D P=0.73) discarding the hypothesis their association with MacTel risk arises from pleiotropic effects of the SNPs used to construct the PRS of these traits.

By comparing SNPs effect size between MacTel disease and the traits we tested for MR association we noticed that the relationship between retinal arteriolar calibre PRS, retinal vascular calibre PRS and retinal thickness PRS with MacTel disease was likely driven by single SNPs in locus 5q14.3 rather than a global effect (Figure S7). To this end, we included SNP rs17421627 in all three models as a covariate. Inclusion of this SNP resulted in retinal arteriolar calibre PRS (b=-0.07, p=0.16) and retinal vascular calibre PRS (b=0.008, p=0.85) to lose significance while macular thickness to change from a positive association a to a negative association (b=-0.088, p=0.046). This last was again found to be entirely driven by SNP rs17279437 in locus 3p21.31 as the inclusion of this SNP resulted in a complete loss of significance between macular thickness and MacTel (b=0.005, p=0.90).
